# Cognitive function in childhood and cardiovascular disease risk factors up to 6 decades later: Pooled data from two birth cohort studies

**DOI:** 10.64898/2026.01.23.26344743

**Authors:** Seyed E Mousavi, Ian J Deary, Steven Bell, Gilad Twig, Mark Hamer, Alun Hughes, G. David Batty

## Abstract

**Background:** Higher childhood (pre-morbid) cognitive function (IQ) appears to confer a lower risk of cardiovascular disease (CVD) episodes in adulthood, however, the underpinning mechanisms are uncertain. We tested the association between childhood cognitive ability and later CVD risk factors that may underpin this gradient.

**Methods:** We used data from two well-characterized prospective birth cohort studies initiated in the United Kingdom in 1958 (N=10870) and 1970 (N=9278). Cognitive function was quantified using validated written tests at around 10 years of age. A total of 16 biological, psychosocial, and behavioral risk factors were collected at 46-48 (1970 cohort) and 61-65 (1958 cohort) years of age in both studies. Associations were summarized using linear and logistic regression.

**Results:** After pooling the data and controlling for early life confounding factors, higher childhood cognitive ability was related to more favorable levels of 4/4 psychosocial, 3/3 behavioral, and 6/9 biological risk factors in middle/older age. The magnitude of these relationships increased from biological, to behavioral then psychosocial risk factors. For example, a one standard deviation higher cognitive test score was related to protective levels of body mass index (beta coefficient [95% confidence interval]: -0.58 [-0.70, -0.45]; p-value <0.001), triglycerides (-0.03 [-0.04, -0.01]; <0.001), cigarette smoking (odds ratio [95% confidence interval]: 0.72 [0.67, 0.76] <0.001), depressive symptoms (0.79 [0.74, 0.84]; <0.001), and a high prestige occupation (2.27 [2.21, 2.35]; <0.001). There was some evidence of mediation via educational attainment.

**Conclusions:** The present correlations implicate a series of mechanistic pathways which may connect cognition with subsequent CVD events. The greater burden of unfavorable risk factors at the lower end of the cognition spectrum suggests that compliance with preventive health advice may be sub-optimal.

**Clinical Perspective What is new?:** This is the most comprehensive examination of cardiovascular disease risk factors in relation to antecedent (pre-morbid) cognitive function (IQ)

Pooling data from two large cohort studies of births from 1958 and 1970 in the UK, children with high cognitive function experienced the most favorable levels of a range of psychosocial, behavioral, and biological CVD risk factors up to six decades later.

**What are the clinical implications?:** These IQ–risk factor correlations implicate a series of mechanistic pathways connecting cognition with future CVD events.

It also seems likely that preventive health advice from clinicians, including lifestyle modification and drug treatments, may be associated with low levels of compliance and/or initiation in people at the lower end of the cognition spectrum.

## Introduction

Cognitive function – also known as mental ability, intelligence, IQ – refers to the capacity to learn, reason, and solve problems.^1^ These characteristics potentially implicate intelligence in the disease prevention process and a growing evidence base has emerged over the last two decades.^2^ Analyses of observational data has revealed seemingly protective effects of high cognitive ability against adverse health outcomes as diverse as dementia,^3^ unintentional^4,5^ and intentional injury,^6^ depression,^7^ digestive disease,^8^ and selected malignancies.^9^ The magnitude of these relationships is typically greater than those apparent for other psychological predictors, including mental illness, occupational stress, neurodevelopmental disorders, and selected personality types.^2^

The most examined health endpoint in this context is cardiovascular disease. In studies of middle-and older-age populations, individuals with higher scores on standard tests of cognitive ability subsequently experienced a lower rate of cardiovascular disease events.^10–12^ Whereas this association does not seem to be explained by socioeconomic background, a key concern has been confounding by comorbid cardiometabolic diseases, such as hypertension^13^ and diabetes,^14^ that in their own right are associated with lower cognitive function and elevate CVD event rates. Recent reports have, however, verified the cognition–CVD gradient in decades-long follow up of general population-based cohorts of children^15^ and young adults^16–19^ in whom the prevalence of such co-morbidities was so low at assessment of cognitive function as to circumvent the problem of confounding. Of note, the risk of CVD occurs in a linear manner across the full childhood IQ range, rather than only being apparent at particular thresholds such as those denoting average intelligence or learning difficulties. Effect sizes for cognitive ability are commensurate with those seen for established CVD risk factors such as high blood pressure and cigarette smoking.^20^

Further, these observations triangulate with those from studies using imaging-based assessment of atherosclerotic load in middle-age^21^ and those examining the genetic correlation between intelligence and CVD.^22^

Despite the consistency of these findings, there remains a paucity of understanding about the mechanisms that might underpin the association of prior cognitive ability with the occurrence of CVD episodes. One obvious possibility is mediation via selected behavioral, psychological, social, and biological risk factors. There is evidence that, by middle-age, children/young people with higher cognitive function tend to have more favorable social characteristics, particularly educational achievement^23^ and other markers of socio-economic status such as income and job prestige.^24^ There are also emerging data that psychological traits such as depressive disorder,^7^ and health behaviors such as cigarette smoking^25–27^ and physical inactivity,^28^ are less prevalent amongst people with higher mental ability. Equivalent evidence for established biological risk factors are, however, currently lacking; indeed, more is understood about the influence of these characteristics on subsequent cognitive performance rather than the converse.^29,30^ The few studies of pre-morbid cognitive ability and later assessment of biological risk factors report marginally lower levels of adult blood pressure^31,32^ and blood cholesterol^33^ in individuals with higher childhood intelligence relative to their lower-scoring counterparts.

In the present study we pooled data from two large UK general population-based birth cohort studies to examine the relationships between antecedent cognitive ability and biological risk factors for CVD in middle/older age, making comparisons of the magnitude of these associations with those apparent for psychosocial and behavioral risk factors.

## Methods

Described in detail elsewhere,^34,35^ the National Child Development Study (hereafter, the 1958 cohort study) (N=18550) and the 1970 cohort study (N=18032) are population-based, ongoing, closed longitudinal studies initiated at the birth of United Kingdom residents. Ethical approval for data collection described herein in the 1958 (reference 12/LO/2010) and 1970 (14/LO/0371) cohorts has been provided by the National Health Service Research Ethics Committee. All participants have given written consent. In composing our manuscript we followed the Strengthening the Reporting of Observational Studies in Epidemiology (STROBE) Statement guidelines.^36^

Tracking the lives of individuals born during a single week in each birth year, there have been 11 phases of data collection in the 1958 cohort and 12 in the 1970 cohort. Pre-adult data collection took place in schools, with data thereafter captured in the home or by mail/telephone interview. Between 2020 and 2024 (age 61-65), members of the 1958 study took part in a home-based medical examination, with corresponding data being collected in the 1970 cohort between 2016 and 2018 (age 46-48). Conducted under the auspices of the Centre for Longitudinal Studies, the protocols are near-identical so facilitating pooling.

### Assessment of cognitive function and confounding factors in childhood

These pre-adult data were based on medical records or questionnaires completed by a midwife, parent, teacher, and, when age permitted, study members. Children were administered cognitive ability tests under examination conditions in schools at age 11 (1958 study) or 10 years (1970 study). In the 1958 cohort, these were the general ability test and the Copying Designs Test.^37^ In the 1970 cohort, mental ability was assessed using a modified version of the British Ability Scales from which scores were derived for word definitions, word similarities, recall of digits, and matrices.^38^ We computed general cognitive ability (termed ‘*g*’) using principal component analysis: in the 1958 study, the first unrotated principal component accounted for 53% of the total variance in test scores, and in the 1970 study it was 58%.

Birth weight (grams) was extracted from medical records. Family (parental) socioeconomic status was based on the occupation of the father using standard schema and categorized into three groups (managerial, technical, or professional; other skilled work; or lower skilled and other work).^39^ Study member psychological distress was measured using the parent-administered Rutter Scale when the children were 7 (1958 study) and 5 years of age (1970 study); ‘caseness’ was denoted by scores exceeding the 80th percentile.^40^

### Assessment of CVD risk factors in adulthood

We used risk factor data collected during nurse-administered interviews, study member self-report, and medical examination at age 62 in the 1958 study and age 46 in the 1970 study. For the purposes of the present analyses, risk factors were arranged thematically: psychosocial, behavioral, and biological.

### Psychosocial risk factors

Highest academic qualification was grouped into four categories (none; high school qualification; Diploma or less; Degree level or higher). Adult occupational social class was based on the same categorization as that described for family social class. Income was reported weekly take-home earnings over the prior year. Psychological distress was measured using the Malaise inventory,^41^ categorized as low (score 0–3) and high (≥4) with the latter denoting caseness. Lastly, adult cognitive function was ascertained in both studies using the same four tests (Delayed Word List Recall; Immediate Word List Recall; Animal Naming Task; and Letter Cancellation).^42^ The first unrotated principal component accounted for 39% of the total variance in both cohorts.

### Behavioral risk factors

Self-reported quantity of alcoholic beverages consumed was converted to units/week (0; ≤14; >14), and cigarette smoking habit based on standard groupings (never; former; current). In the 1958 study, physical activity was denoted by the number of occasions exertion was vigorous enough to produce a sweat response and categorized based on frequency, whereas in the 1970 study enquiries were made about the frequency of 30-minute bouts.^43^ The same classification was used in both studies (0; 1–3; ≥4 days/week). In the 1970 study only, physical activity was measured objectively using a thigh-worn accelerometer,^44^ yielding data on daily duration of moderate/vigorous exertion (hours/day), daily step count, and total daily activity (hours/day).

Dietary intake was quantified on-line using the Oxford WebQ Questionnaire^45^ with dietary fiber intake and the ratio of saturated fat : total fat then derived.

### Biological risk factors

Following a 5-minute rest, blood pressure and resting heart rate was measured twice (1958 study) or thrice (1970 study) in a seated position using the Omron HEM 907 monitor and the average value used in our analyses. For study members taking blood pressure-lowering medications, readings were corrected by adding 10 mmHg.^46^ As expected, with correlation coefficients between systolic and diastolic blood pressure in both the 1958 (0.72, p-value<0.001) and 1970 cohorts being high (0.83, p-value<0.001), we show data for systolic blood pressure only. Height was measured directly using a stadiometer and weight using Tanita BF-522W scales. Body mass index (BMI) was computed using standard formulae.^47^ Following blood draw, serum total cholesterol, high-density lipoprotein (HDL) cholesterol, and triglycerides were assayed using enzymatic colorimetric methods with correction made for lipid-regulating medication (+25% for total cholesterol; −6% for HDL cholesterol; and +18% for triglycerides).^48^ High-sensitivity C-reactive protein was assessed using particle-enhanced immunoturbidimetry, and glycated hemoglobin (HbA1c) was measured using ion-exchange high-performance liquid chromatography. HbA1c values were adjusted (+11 mmol/mol) for participants taking medication for blood glucose control and a percentage conversion made using the standard formulae.^49^

### Statistical analyses

To examine potential confounder and risk-factor levels according to childhood cognitive function, we used a one-way ANOVA test for parametric continuous variables, the Kruskal–Wallis rank-sum test for non-parametric continuous variables, and the Chi-square test for categorical variables.

General cognitive ability was standardized (mean 100, standard deviation 15) for use in regression models. Skewed outcome data – triglycerides, HbA1C, C-reactive protein, income – were natural log-transformed.

Quartiles of cognitive test scores are used for illustration of differences in adult risk factor levels and continua utilized in regression analyses (linear, logistic, and multinomial). With preliminary regression analyses revealing no convincing evidence of effect modification by sex, within-cohort effects estimates were pooled and sex-adjusted. Being birth cohorts born in single weeks in their respective birth years, no statistical adjustment for age was required.

In study-specific analyses of cognition and later CVD risk factors, effect estimates were initially adjusted for sex (model 1) and then sex plus other potential confounders (parental social class, birthweight, and psychological distress in early life) (model 2). Given the known interrelationship between childhood cognition and later educational attainment,^23^ including in the present studies (r=0.47, p-value<0.001 in the 1958 cohort; r=0.40, p-value< 0.001 in the 1970 cohort), we conceptualized education as a mediator and scrutinized its impact by separately adding it to model 2. Study-specific effect estimates were pooled using fixed-effect inverse-variance meta-analysis. All statistical analyses were conducted using R version 4.3.3 (Vienna, Austria).

## Results

In the 1958 cohort, 10870 people (5329 women) comprised the analytical sample and in the 1970 study there were 9278 individuals (4490 women) (figure 1). We compared the early life (pre-attrition) characteristics of these individuals with those omitted and differences were generally modest, suggesting little evidence of selection bias (supplemental tables 1 and 2). There were, however, marked variations in potential confounding factors according to cognition in both studies, whereby the most favorable levels were consistently apparent in children who performed better on the ability tests. In the 1958 study for instance, relative to children in the lowest cognition group, those in the highest were, on average, more than four times more likely to have a father working in a higher prestige profession (30 versus 7%).

**Figure 1.**
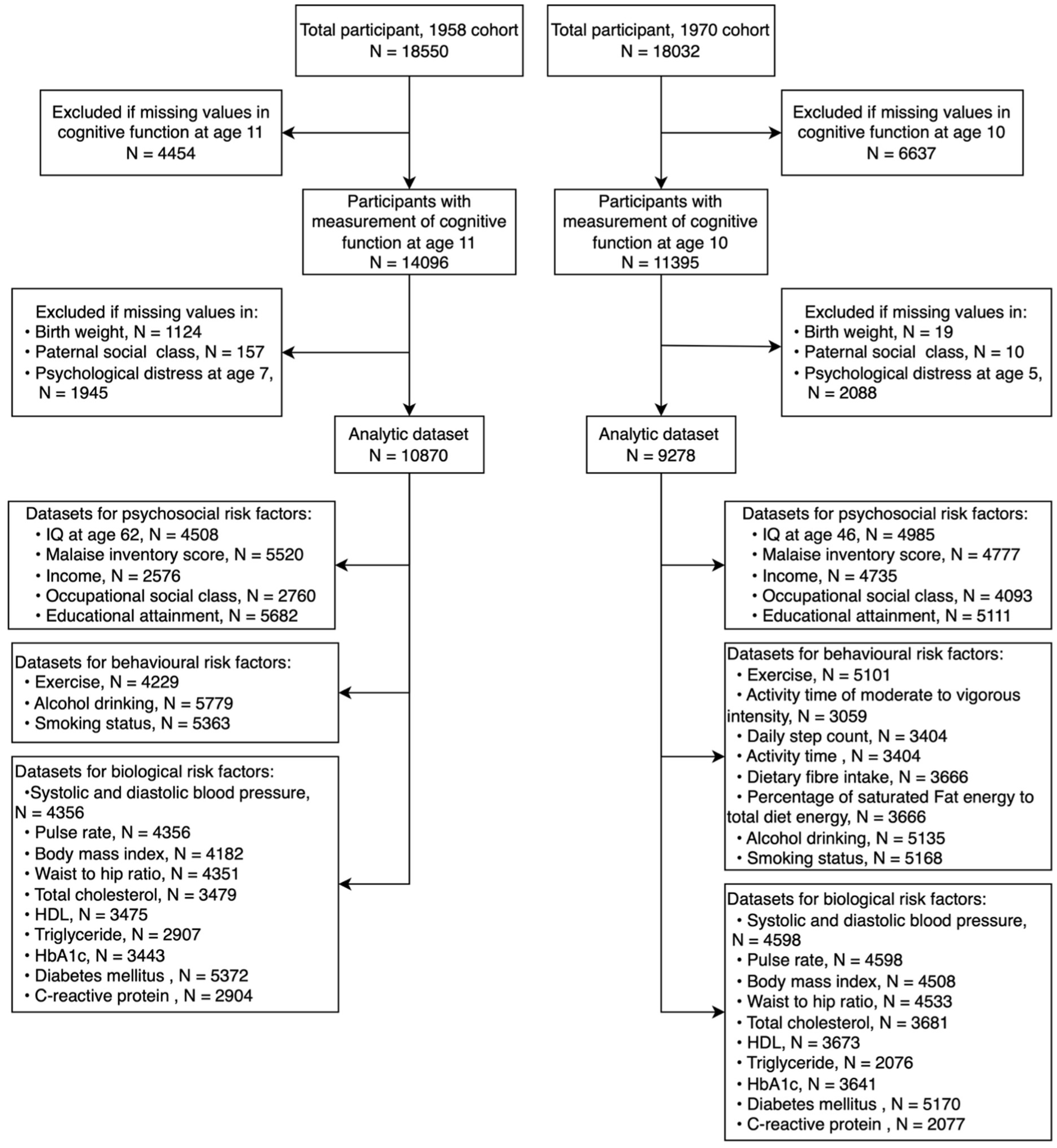
Derivation of analytic samples: 1958 and 1970 birth cohort studies

Next, we examined differences in psychosocial, behavioral, and biological risk factors in middle-(1970 study) and older-age (1958 study) according to categories of childhood cognition. After pooling the data and controlling for early life confounding factors, higher childhood cognitive ability was related to more favorable levels of 4/4 psychosocial, 3/3 behavioral, and 6/9 biological risk factors in middle/older age, as follows.

### Psychosocial risk factors in adulthood

Across quartiles of childhood IQ groups, there were stark differentials in indices of adult socioeconomic status, including educational achievement, occupational social class, and income (supplemental tables 3 and 4). Symptoms of psychological distress were also less prevalent in adults who had scored more highly on cognition tests in childhood. Stepwise effects were typically apparent. In figures 2 (continuous outcome variables) and 3 (categorical) we summarize the results from regression models for these relationships. For adult socio-economic outcomes in both studies there were clear and consistent associations with earlier IQ whereby, in confounder-adjusted analyses of the pooled data, a one standard deviation higher cognition was associated with a 2-fold greater likelihood of being in a managerial occupation in later life (odds ratio; 95% confidence interval: 2.27; 2.21, 2.35), a 5-fold greater likelihood of receiving a university degree (4.70; 4.51, 4.91), and having a higher income (beta coefficient; 95% confidence interval: 0.15; 0.13, 0.18). Mental health was also more favorable in the higher cognitive test scorers such that a lower risk of psychological distress un later life was apparent (0.79; 0.74, 0.84). There was some attenuation seen in the magnitude of effect estimates after adult educational attainment was added to the multivariable model, potentially implicating this characteristic as a partial mediator in some of these associations (table 1).

**Figure 2.**
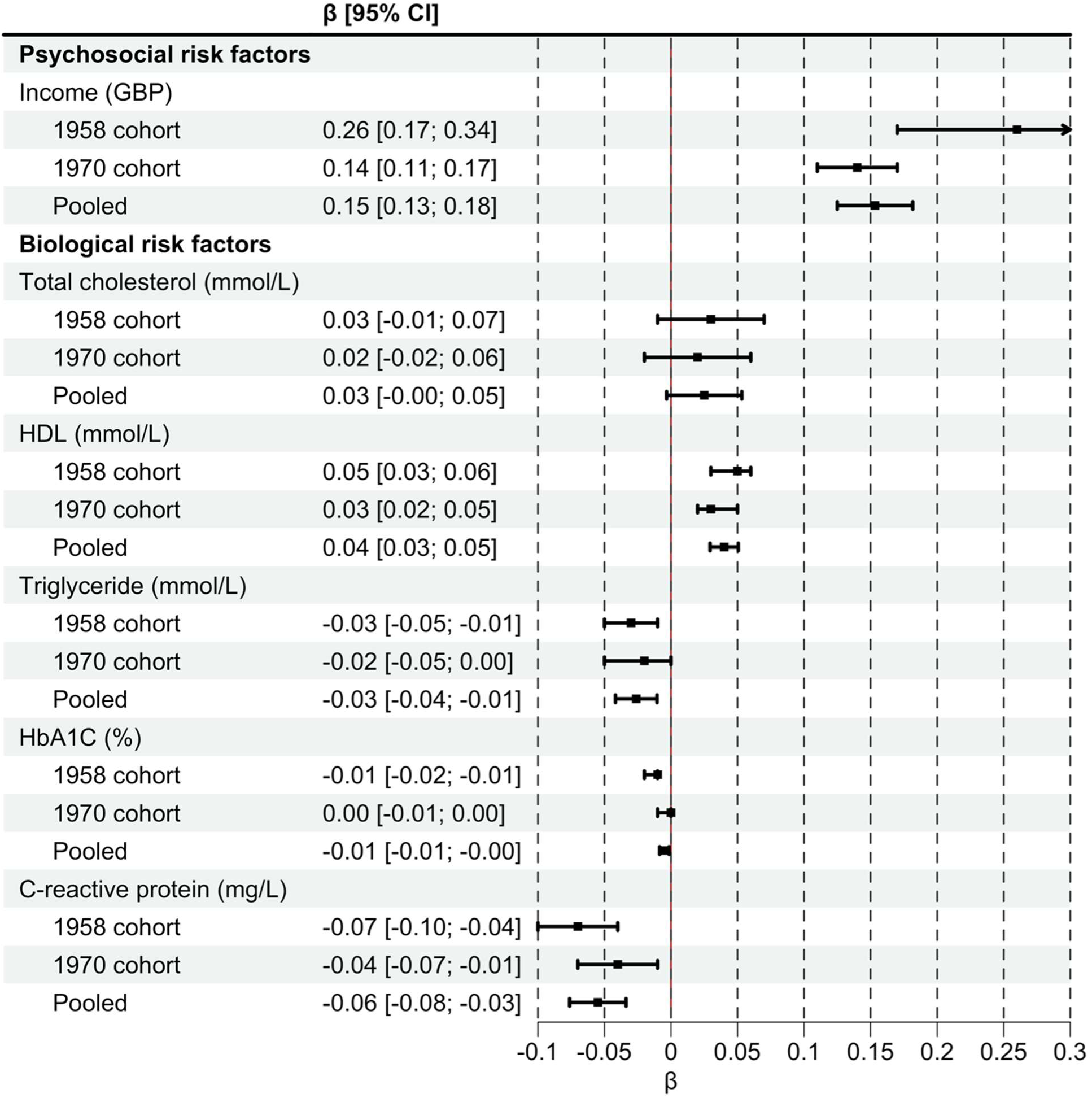
Beta coefficients (95% confidence intervals) for a one standard deviation higher childhood IQ with cardiovascular disease risk factors in the 1958 (age 62) and 1970 (age 46) cohorts

**Figure 3.**
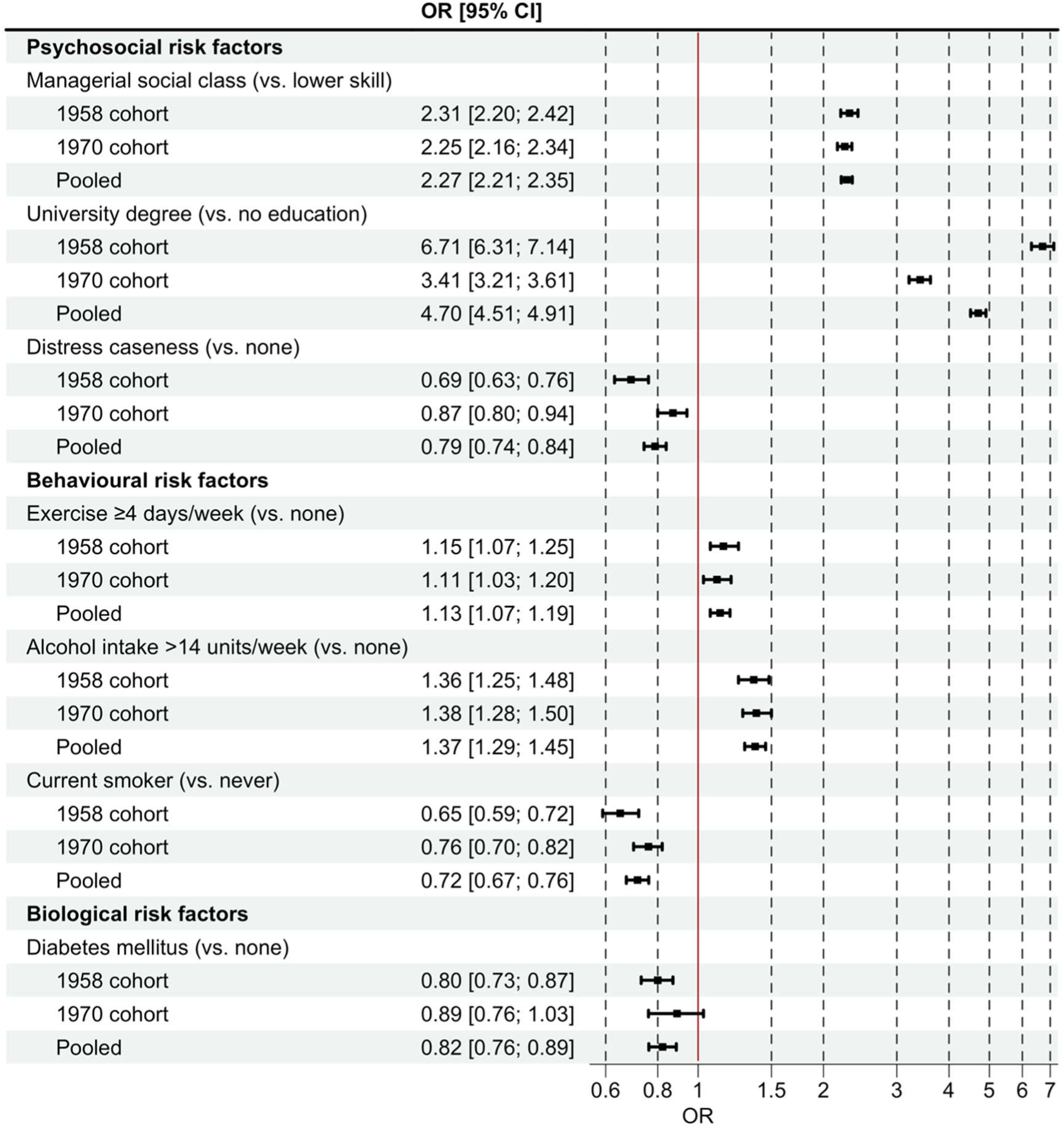
Odds ratio (95% confidence interval) for a one standard deviation higher childhood IQ with cardiovascular disease risk factors in the 1958 (age 62) and 1970 (age 46) cohorts

**Table 1.**
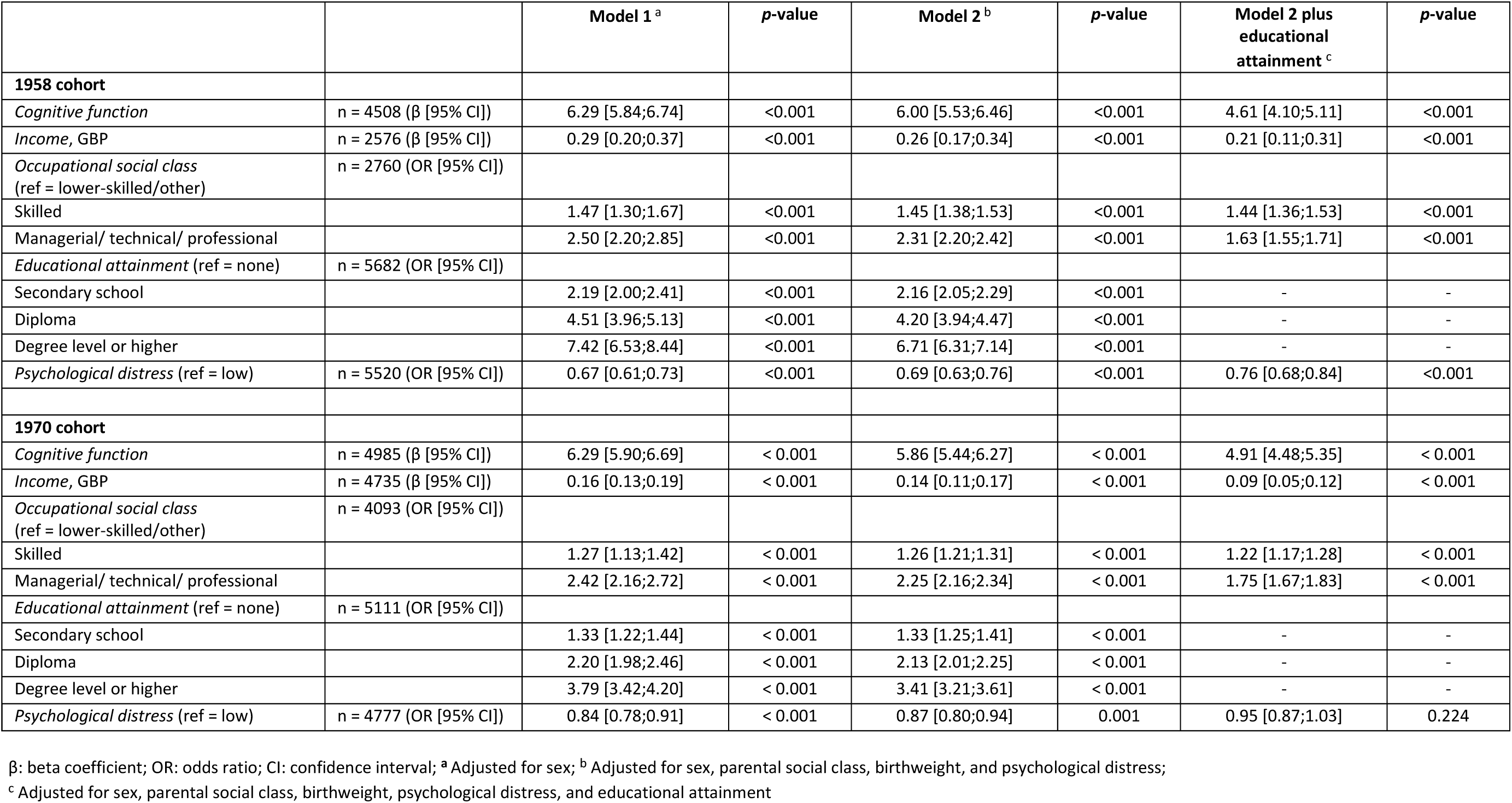
Multivariable associations for a one standard deviation higher childhood IQ with psychosocial risk factors in the 1958 (age 62) and 1970 (age 46) cohort studies

|  |  | Model 1 <sup>a</sup> | p-value | Model 2 <sup>b</sup> | p-value | Model 2 plus educational attainment <sup>c</sup> | p-value |
| --- | --- | --- | --- | --- | --- | --- | --- |
| **1958 cohort** |  |  |  |  |  |  |  |
| *Cognitive function* | n = 4508 (β [95% CI]) | 6.29 [5.84;6.74] | <0.001 | 6.00 [5.53;6.46] | <0.001 | 4.61 [4.10;5.11] | <0.001 |
| *Income, GBP* | n = 2576 (β [95% CI]) | 0.29 [0.20;0.37] | <0.001 | 0.26 [0.17;0.34] | <0.001 | 0.21 [0.11;0.31] | <0.001 |
| *Occupational social class*<br>(ref = lower-skilled/other) | n = 2760 (OR [95% CI]) |  |  |  |  |  |  |
| Skilled |  | 1.47 [1.30;1.67] | <0.001 | 1.45 [1.38;1.53] | <0.001 | 1.44 [1.36;1.53] | <0.001 |
| Managerial/ technical/ professional |  | 2.50 [2.20;2.85] | <0.001 | 2.31 [2.20;2.42] | <0.001 | 1.63 [1.55;1.71] | <0.001 |
| *Educational attainment* (ref = none) | n = 5682 (OR [95% CI]) |  |  |  |  |  |  |
| Secondary school |  | 2.19 [2.00;2.41] | <0.001 | 2.16 [2.05;2.29] | <0.001 | - | - |
| Diploma |  | 4.51 [3.96;5.13] | <0.001 | 4.20 [3.94;4.47] | <0.001 | - | - |
| Degree level or higher |  | 7.42 [6.53;8.44] | <0.001 | 6.71 [6.31;7.14] | <0.001 | - | - |
| *Psychological distress* (ref = low) | n = 5520 (OR [95% CI]) | 0.67 [0.61;0.73] | <0.001 | 0.69 [0.63;0.76] | <0.001 | 0.76 [0.68;0.84] | <0.001 |
| **1970 cohort** |  |  |  |  |  |  |  |
| *Cognitive function* | n = 4985 (β [95% CI]) | 6.29 [5.90;6.69] | < 0.001 | 5.86 [5.44;6.27] | < 0.001 | 4.91 [4.48;5.35] | < 0.001 |
| *Income, GBP* | n = 4735 (β [95% CI]) | 0.16 [0.13;0.19] | < 0.001 | 0.14 [0.11;0.17] | < 0.001 | 0.09 [0.05;0.12] | < 0.001 |
| *Occupational social class*<br>(ref = lower-skilled/other) | n = 4093 (OR [95% CI]) |  |  |  |  |  |  |
| Skilled |  | 1.27 [1.13;1.42] | < 0.001 | 1.26 [1.21;1.31] | < 0.001 | 1.22 [1.17;1.28] | < 0.001 |
| Managerial/ technical/ professional |  | 2.42 [2.16;2.72] | < 0.001 | 2.25 [2.16;2.34] | < 0.001 | 1.75 [1.67;1.83] | < 0.001 |
| *Educational attainment* (ref = none) | n = 5111 (OR [95% CI]) |  |  |  |  |  |  |
| Secondary school |  | 1.33 [1.22;1.44] | < 0.001 | 1.33 [1.25;1.41] | < 0.001 | - | - |
| Diploma |  | 2.20 [1.98;2.46] | < 0.001 | 2.13 [2.01;2.25] | < 0.001 | - | - |
| Degree level or higher |  | 3.79 [3.42;4.20] | < 0.001 | 3.41 [3.21;3.61] | < 0.001 | - | - |
| *Psychological distress* (ref = low) | n = 4777 (OR [95% CI]) | 0.84 [0.78;0.91] | < 0.001 | 0.87 [0.80;0.94] | 0.001 | 0.95 [0.87;1.03] | 0.224 |

β: beta coefficient; OR: odds ratio; CI: confidence interval; <sup>a</sup> Adjusted for sex; <sup>b</sup> Adjusted for sex, parental social class, birthweight, and psychological distress;
<sup>c</sup> Adjusted for sex, parental social class, birthweight, psychological distress, and educational attainment

### Behavioral risk factors in adulthood

In both cohorts, children with higher mental ability were typically more likely to choose healthy behaviors as adults (supplemental tables 5 and 6). As such, those with higher intelligence test scores, on average, avoided regular smoking; were more physically active in middle/older age, particularly with moderate frequency, although there were no clear differences in various markers of accelerometry-captured physical exertion (1970 cohort only, supplemental table 6); and had a marginally higher dietary fiber intake. Again, many of these effects were stepwise across the full cognition range. A clear departure from this pattern of cardiovascular protection in the higher IQ groups was a greater total intake of alcohol in later life in study members with high pre-adult cognitive function.

In multivariable analyses, the magnitude of the relationships between cognition and health behaviors were little attenuated after adjustment for early life covariates (figures 2 and 3). After multiple confounder adjustment, in the pooled dataset a one standard higher cognition score was associated with a 28% lower likelihood of being a cigarette smoker (0.72; 0.67, 0.76), a 13% greater probability of exercising on 1 to 3 occasions/week (1.13; 1.07, 1.19), and 37% greater chance of drinking more than 14 units of alcohol/week (1.37; 1.29, 1.45). Taking adult education into account again impacted the point estimates although statistical significance at conventional levels was largely retained (table 2).

**Table 2.** Multivariable associations for a one standard deviation higher childhood IQ with behavioural risk factors in the 1958 (age 62) and 1970 (age 46) cohort studies

|  |  | Model 1 <sup>a</sup> | p-value | Model 2 <sup>b</sup> | p-value | Model 2 plus educational attainment <sup>c</sup> | p-value |
| --- | --- | --- | --- | --- | --- | --- | --- |
| **1958 cohort** |  |  |  |  |  |  |  |
| *Alcohol intake, units/week (ref = none)* | n = 5779 (OR [95% CI]) |  |  |  |  |  |  |
| ≤ 14 |  | 1.39 [1.30;1.48] | <0.001 | 1.35 [1.26;1.44] | <0.001 | 1.23 [1.15;1.31] | <0.001 |
| > 14 |  | 1.42 [1.31;1.55] | <0.001 | 1.36 [1.25;1.48] | <0.001 | 1.31 [1.20;1.41] | <0.001 |
| *Cigarette smoking (ref = never smoked)* | n = 5363 (OR [95% CI]) |  |  |  |  |  |  |
| Former smoker |  | 0.84 [0.79;0.90] | <0.001 | 0.85 [0.80;0.91] | <0.001 | 0.95 [0.89;1.01] | 0.117 |
| Current smoker |  | 0.63 [0.57;0.69] | <0.001 | 0.65 [0.59;0.72] | <0.001 | 0.84 [0.76;0.92] | <0.001 |
| *Exercise, days/week (ref = none)* | n = 4229 (OR [95% CI]) |  |  |  |  |  |  |
| 1-3 |  | 1.44 [1.32;1.56] | <0.001 | 1.38 [1.27;1.50] | <0.001 | 1.22 [1.13;1.33] | <0.001 |
| ≥ 4 |  | 1.19 [1.10;1.29] | <0.001 | 1.15 [1.07;1.25] | <0.001 | 1.05 [0.97;1.14] | 0.198 |
| **1970 cohort** |  |  |  |  |  |  |  |
| *Alcohol intake, units/week (ref = none)* | n = 5135 (OR [95% CI]) |  |  |  |  |  |  |
| ≤ 14 |  | 1.39 [1.29;1.49] | < 0.001 | 1.33 [1.24;1.43] | < 0.001 | 1.20 [1.12;1.29] | < 0.001 |
| > 14 |  | 1.45 [1.34;1.57] | < 0.001 | 1.38 [1.28;1.50] | < 0.001 | 1.33 [1.23;1.44] | < 0.001 |
| *Cigarette smoking (ref = never smoked)* | n = 5168 (OR [95% CI]) |  |  |  |  |  |  |
| Former smoker |  | 0.99 [0.93;1.06] | 0.812 | 0.99 [0.93;1.06] | 0.722 | 1.11 [1.04;1.19] | 0.002 |
| Current smoker |  | 0.73 [0.68;0.79] | < 0.001 | 0.76 [0.70;0.82] | < 0.001 | 0.92 [0.85;1.00] | 0.039 |
| *Exercise, days/week (ref = none)* | n = 5101 (OR [95% CI]) |  |  |  |  |  |  |
| 1-3 |  | 1.52 [1.40;1.64] | < 0.001 | 1.45 [1.34;1.56] | < 0.001 | 1.28 [1.18;1.38] | < 0.001 |
| ≥ 4 |  | 1.15 [1.06;1.24] | < 0.001 | 1.11 [1.03;1.20] | 0.007 | 1.10 [1.02;1.18] | 0.010 |
| *MVI activity, hours/day* | n = 3059 (β [95% CI]) | 0.01 [-0.01;0.03] | 0.200 | 0.00 [-0.01;0.02] | 0.805 | -0.01 [-0.03;0.01] | 0.437 |
| *Mean daily step count* | n = 3404 (β [95% CI]) | 8.04 [-120.02;136.10] | 0.902 | -66.27 [-200.58;68.05] | 0.333 | -107.41 [-252.60;37.79] | 0.147 |
| *Mean daily activity, hours* | n = 3404 (β [95% CI]) | -0.03 [-0.05;0.00] | 0.023 | -0.04 [-0.07;-0.01] | 0.003 | -0.04 [-0.07;-0.01] | 0.010 |
| *Dietary fibre intake, gram* | n = 3666 (β [95% CI]) | 0.40 [0.16;0.64] | 0.001 | 0.26 [0.01;0.52] | 0.044 | -0.04 [-0.31;0.23] | 0.752 |
| *Saturated fat : total fat, percent* | n = 3666 (β [95% CI]) | 0.06 [-0.06;0.18] | 0.353 | 0.07 [-0.06;0.20] | 0.285 | 0.08 [-0.05;0.22] | 0.237 |

β: beta coefficient; OR: odds ratio; CI: confidence interval. <sup>a</sup> Adjusted for sex; <sup>b</sup> Adjusted for sex, parental social class, birthweight, and psychological distress;
<sup>c</sup> Adjusted for sex, parental social class, birthweight, psychological distress, and educational attainment; MVI, moderate-to-vigorous intensity

### Biological risk factors in adulthood

Of the biological risk factors ascertained in later life, in both studies there was a suggestion of more favorable levels of systolic blood pressure, resting heart rate, body weight, HDL cholesterol, triglycerides, HbA1C, and systemic inflammation in middle- and older-age adults who performed better on antecedent cognitive tests (supplemental tables 7 and 8). In general, levels of these risk factors were monotonic across the full IQ gradient, however, while statistical significance at conventional levels was commonplace, absolute differences across cognition groups were often modest and lower in magnitude than those seen for the psychosocial and behavioral risk factors. After pooling data from both studies and adjusting for confounding factors, higher pre-adult cognitive function was associated with more favorable levels of BMI (beta coefficient; 95% confidence interval: -0.58; -0.70, -0.45), waist-to-hip ratio (-0.89; -1.05, -0.73), resting heart rate (-0.55; -0.81, -0.30), HDL (0.04; 0.03, 0.05), triglycerides (-0.03; -0.04, -0.01), and C-reactive protein (-0.06; -0.08, -0.03) (table 3 and figure 2). While HbA1C was not associated with earlier measurement of cognitive function after statistical control (-0.01; -0.01, -0.00), higher mental ability nonetheless appeared to confer protection against type 2 diabetes (odds ratio; 95% confidence interval: 0.82; 0.76, 0.89). Again, controlling for educational achievement led to attenuation in the strength of these relationships with loss of statistical significance for selected outcomes.

**Table 3.**
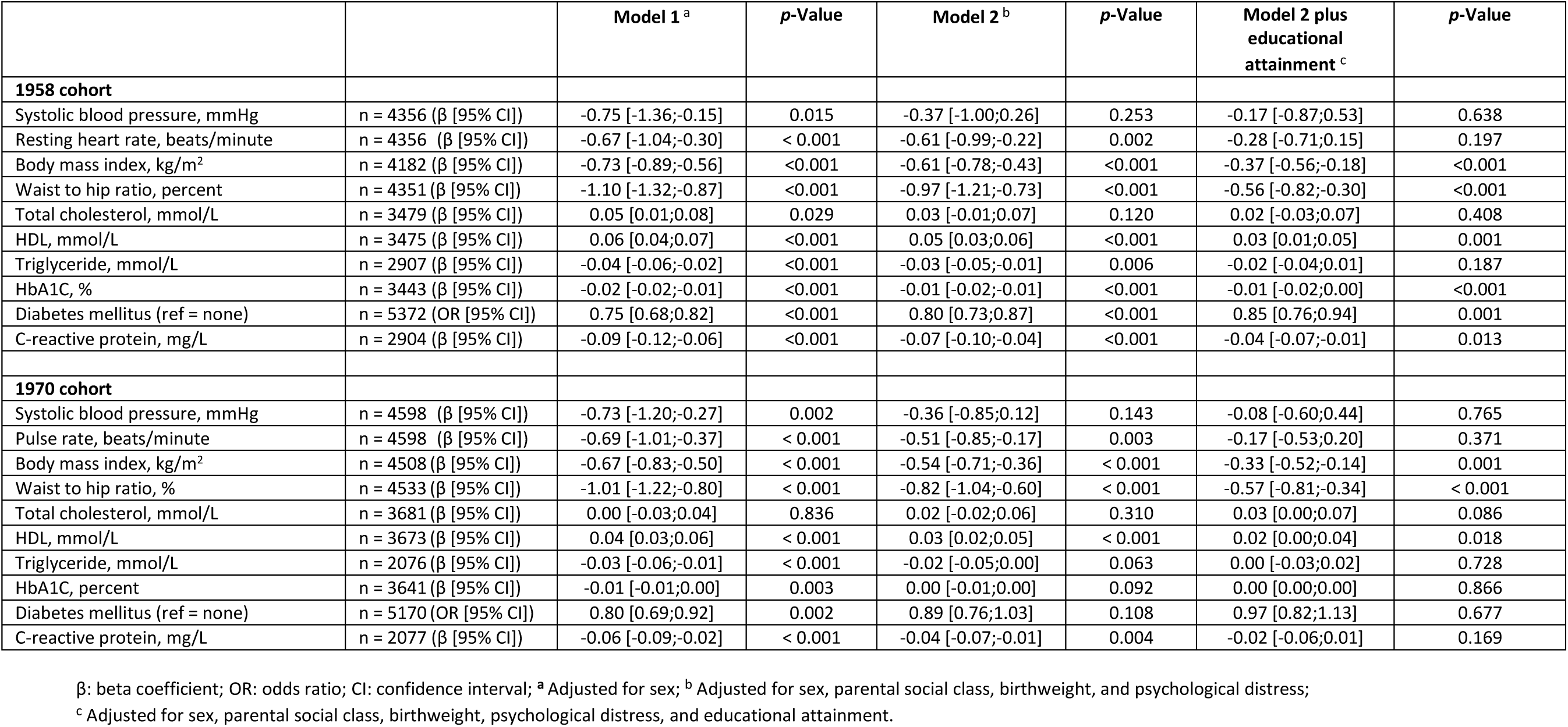
Multivariable associations for a one standard deviation higher childhood IQ with biological risk factors in the 1958 (age 62) and 1970 (age 46) cohort studies

|  |  | Model 1 <sup>a</sup> | p-Value | Model 2 <sup>b</sup> | p-Value | Model 2 plus educational attainment <sup>c</sup> | p-Value |
| --- | --- | --- | --- | --- | --- | --- | --- |
| **1958 cohort** |  |  |  |  |  |  |  |
| Systolic blood pressure, mmHg | n = 4356 (β [95% CI]) | -0.75 [-1.36;-0.15] | 0.015 | -0.37 [-1.00;0.26] | 0.253 | -0.17 [-0.87;0.53] | 0.638 |
| Resting heart rate, beats/minute | n = 4356 (β [95% CI]) | -0.67 [-1.04;-0.30] | < 0.001 | -0.61 [-0.99;-0.22] | 0.002 | -0.28 [-0.71;0.15] | 0.197 |
| Body mass index, kg/m<sup>2</sup> | n = 4182 (β [95% CI]) | -0.73 [-0.89;-0.56] | <0.001 | -0.61 [-0.78;-0.43] | <0.001 | -0.37 [-0.56;-0.18] | <0.001 |
| Waist to hip ratio, percent | n = 4351 (β [95% CI]) | -1.10 [-1.32;-0.87] | <0.001 | -0.97 [-1.21;-0.73] | <0.001 | -0.56 [-0.82;-0.30] | <0.001 |
| Total cholesterol, mmol/L | n = 3479 (β [95% CI]) | 0.05 [0.01;0.08] | 0.029 | 0.03 [-0.01;0.07] | 0.120 | 0.02 [-0.03;0.07] | 0.408 |
| HDL, mmol/L | n = 3475 (β [95% CI]) | 0.06 [0.04;0.07] | <0.001 | 0.05 [0.03;0.06] | <0.001 | 0.03 [0.01;0.05] | 0.001 |
| Triglyceride, mmol/L | n = 2907 (β [95% CI]) | -0.04 [-0.06;-0.02] | <0.001 | -0.03 [-0.05;-0.01] | 0.006 | -0.02 [-0.04;0.01] | 0.187 |
| HbA1C, % | n = 3443 (β [95% CI]) | -0.02 [-0.02;-0.01] | <0.001 | -0.01 [-0.02;-0.01] | <0.001 | -0.01 [-0.02;0.00] | <0.001 |
| Diabetes mellitus (ref = none) | n = 5372 (OR [95% CI]) | 0.75 [0.68;0.82] | <0.001 | 0.80 [0.73;0.87] | <0.001 | 0.85 [0.76;0.94] | 0.001 |
| C-reactive protein, mg/L | n = 2904 (β [95% CI]) | -0.09 [-0.12;-0.06] | <0.001 | -0.07 [-0.10;-0.04] | <0.001 | -0.04 [-0.07;-0.01] | 0.013 |
| **1970 cohort** |  |  |  |  |  |  |  |
| Systolic blood pressure, mmHg | n = 4598 (β [95% CI]) | -0.73 [-1.20;-0.27] | 0.002 | -0.36 [-0.85;0.12] | 0.143 | -0.08 [-0.60;0.44] | 0.765 |
| Pulse rate, beats/minute | n = 4598 (β [95% CI]) | -0.69 [-1.01;-0.37] | < 0.001 | -0.51 [-0.85;-0.17] | 0.003 | -0.17 [-0.53;0.20] | 0.371 |
| Body mass index, kg/m<sup>2</sup> | n = 4508 (β [95% CI]) | -0.67 [-0.83;-0.50] | < 0.001 | -0.54 [-0.71;-0.36] | < 0.001 | -0.33 [-0.52;-0.14] | 0.001 |
| Waist to hip ratio, % | n = 4533 (β [95% CI]) | -1.01 [-1.22;-0.80] | < 0.001 | -0.82 [-1.04;-0.60] | < 0.001 | -0.57 [-0.81;-0.34] | < 0.001 |
| Total cholesterol, mmol/L | n = 3681 (β [95% CI]) | 0.00 [-0.03;0.04] | 0.836 | 0.02 [-0.02;0.06] | 0.310 | 0.03 [0.00;0.07] | 0.086 |
| HDL, mmol/L | n = 3673 (β [95% CI]) | 0.04 [0.03;0.06] | < 0.001 | 0.03 [0.02;0.05] | < 0.001 | 0.02 [0.00;0.04] | 0.018 |
| Triglyceride, mmol/L | n = 2076 (β [95% CI]) | -0.03 [-0.06;-0.01] | < 0.001 | -0.02 [-0.05;0.00] | 0.063 | 0.00 [-0.03;0.02] | 0.728 |
| HbA1C, percent | n = 3641 (β [95% CI]) | -0.01 [-0.01;0.00] | 0.003 | 0.00 [-0.01;0.00] | 0.092 | 0.00 [0.00;0.00] | 0.866 |
| Diabetes mellitus (ref = none) | n = 5170 (OR [95% CI]) | 0.80 [0.69;0.92] | 0.002 | 0.89 [0.76;1.03] | 0.108 | 0.97 [0.82;1.13] | 0.677 |
| C-reactive protein, mg/L | n = 2077 (β [95% CI]) | -0.06 [-0.09;-0.02] | < 0.001 | -0.04 [-0.07;-0.01] | 0.004 | -0.02 [-0.06;0.01] | 0.169 |

β: beta coefficient; OR: odds ratio; CI: confidence interval; <sup>a</sup> Adjusted for sex; <sup>b</sup> Adjusted for sex, parental social class, birthweight, and psychological distress;
<sup>c</sup> Adjusted for sex, parental social class, birthweight, psychological distress, and educational attainment.

## Discussion

In the most comprehensive examination of the connection between early life cognition and later measurement of CVD risk factors, we found that IQ was related to a protective pattern of vascular risk factors in middle- and older-age people in two national birth cohort studies. Associations of ascending magnitude were found for biological, followed by behavioral, and psychosocial risk factors.

Throughout our analyses, controlling for educational attainment consistently lowered the magnitude of the effects estimates and statistical significance at conventional levels was lost on occasion, suggesting some mediating influence. Given the moderate-to-high correlation with IQ, our exposure of interest, the inclusion of education in the multivariable models is moot. As we have elsewhere,^26,50,51^ in the interest of transparency, we computed effect estimates pre- and post-adjustment for education.

### Comparison with existing studies

While there is growing evidence that children/young people with higher cognitive function tend have more favorable socioeconomic trajectories – they typically perform better educationally^23^ alongside having a higher income and occupational status^24^ – as described, there is less clarity as to the potential impact of mental ability on subsequently assessed health behaviors and biomarkers. Cigarette smoking^25–27^ and physical inactivity^28,52^ appear to be less commonplace in adults who, as children and adolescents, had higher scores on standard test of cognitive function.

Our observation of lower resting heart rate – a marker of cardiorespiratory fitness^53^ – in people with higher antecedent IQ accords with these results for self-reported physical activity, including those herein, though we found no relationship with accelerometry-derived indices of this behavior.

That greater total alcohol consumption was apparent in people with higher cognitive function may seem to run counter to the results for other health behaviors, however, taking the evidence in aggregate, there is an explicable differential effect according to the outcome of interest: total consumed versus pattern of consumption. Moderate total alcohol intake, which was widely regarded as health-giving during the period of data collection herein, is typically higher in people with high IQ scores.^52,54^ By contrast, binge drinking, which is known to be cardio-toxic,^55^ is markedly less common in adults who, as children, had better performance on cognitive tests. This is evident whether the habit is defined by a high number of drinks consumed in a single sitting^52,56^ or the physiological consequence, hangovers.^51^

With a higher level of physical exertion apparent in higher IQ scoring people in the present study, it would be anticipated that this group would also be leaner and, of the biological risk factors, cognition was most consistently related to lower body weight and waste-to-hip ratio. Systematic reviews of the observational data suggest this is a universal finding.^57^ It would follow that levels of other closely connected biological risk factors would reveal similar associations with antecedent IQ. While this was the case for triglycerides and total and HDL cholesterol, after confounder-adjustments, we found no association with blood pressure nor glycated hemoglobin. Diabetes was, however, less prevalent in people with higher cognition scores, an association reported elsewhere.^58^ The few existing studies of pre-morbid cognitive ability and later biological risk indices tend to report marginally lower levels of adult blood pressure^31,32^ and blood cholesterol.^33^

### Study strengths and weaknesses

Our report has several strengths including cross-study and cross-risk factor comparisons using population-based data which offer generalizability. Inevitably, our work also has its shortcomings. First, we did not have data on hospitalizations and/or deaths ascribed to CVD to formally test if the risk factors associated with cognition lay on the causal pathway. Second, cognition was ascertained using standard written examinations of intelligence rather than measured objectively using, for instance, stimulus-triggered reaction time which, as well as being strongly inversely correlated with cognitive function, also reveals consistent associations with vascular events.^59^ Third, we did not collect data on some emerging risk factors such as interleukin-6, a marker of systemic inflammation causally linked to CVD^60^ nor biomarkers of vitamin consumption.^61^ Fourth, we carried out the standard correction for blood pressure and HbA1C in study members who reported drug therapy. People with modest educational attainment – a close correlate of cognitive function – are less likely to adhere to drug treatment for raised blood pressure.^62^ This may have led to some overcorrection for blood pressure levels with the potential for underestimation of the true relationship with IQ. Lastly, as an observational study, our results are subject to the perennial concerns of confounding, both residual and unmeasured. A potential solution would be extended follow up of participants in randomized controlled trials which produced cognitive advantage amongst the participants in the intervention group (e.g., the Head Start programme^63^), though this would be logistically challenging.

### Research and clinical implications

The correlation between an array of adult risk factors and antecedent cognition implicates a series of mechanistic pathways which connect cognition with future CVD events. We have provided grounds for formal testing of this hypothesis. On a policy level, a greater prevalence of unfavorable behavioral and biological risk factor levels at the lower end of the cognition spectrum suggests that existing preventive health advice from clinicians – lifestyle modification and drug treatments – may be overly complex for some people. Some recalibration of messaging and delivery may therefore be indicated.

In conclusion, in the most comprehensive analysis of which we are aware, in a pooling of two large birth cohort studies, children with high cognitive function experienced the most favorable levels of psychosocial, behavioral, and biological risk factors up to six decades later. Some of these factors, such as diabetes, may both lie on the mechanistic pathway linking early life cognition with CVD event risk and be important adverse endpoints in their own right.

## Data Availability

Data from both studies are freely available to download via the UK Data Service (https://ukdataservice.ac.uk/).

## Contributions

GDB generated the idea for the present manuscript, developed an analytical plan, and wrote the manuscript. IJD, SB, MH, and AH commented on a draft of the manuscript. SEM built the datasets, developed an analytical plan, conducted the data analyses, and commented on a draft of the manuscript.

